# A Development and Implementation of a Preconception Counseling Program for Black Women and Men in the Southeastern United States: A Pilot Protocol

**DOI:** 10.1101/2024.04.22.24306171

**Authors:** N. Hernandez-Green, M. Haiman, A. McDonald, L. Rollins, C.G. Franklin, O.T.O Farinu, L. Clarke, A. Huebshmann, M. Fort, R. Chandler, P. Brocke, D. McLaurin-Glass, E. Harris, K. Berry, A. Suarez, T. Williams

## Abstract

**Introduction:** Racial/ethnic disparities in maternal mortality rates represent one of the most significant areas of disparities amongst all conventional population perinatal health measures in the U.S. The alarming trends and persistent disparities of outcomes by race/ethnicity and geographic location reinforce the need to focus on ensuring quality and safety of maternity care for all women. Despite complex multilevel factors impacting maternal mortality and morbidity, there are evidence-based interventions that, when facilitated consistently and properly, are known to improve the health of mothers before, during and after pregnancy. The objective of this project is to test implementation of pre-conception counseling with father involvement in community-based settings to improve cardiovascular health outcomes before and during pregnancy in southeastern United States.

**Methods and Analysis:** This study has two components: a comprehensive needs and assets assessment and a small-scale pilot study. We will conduct a community informed needs and assets assessment with our diverse stakeholders to identify opportunities and barriers to preconception counseling as well as develop a stakeholder-informed implementation plan. Next, we will use the implementation plan to pilot preconception counseling with father involvement in community-based settings. Finally, we will critically assess the context, identify potential barriers and facilitators, and iteratively adapt the way preconception counseling can be implemented in diverse settings. Results of this research will support future research focused on identifying barriers and opportunities for scalable and sustainable public health approaches to implementing evidence-based strategies that reduce maternal morbidity and mortality in the southeastern United States’ vulnerable communities.

**Discussion:** Findings will demonstrate that preconception counseling can be implemented in community health settings in the southeastern United States. Furthermore, this study will build the capacity of community-based organizations in addressing the preconception health of their clients. We plan for this pilot to inform a larger scaled-up clinical trial across community health settings in multiple southeastern states.

## 1 Introduction

Racial/ethnic disparities in maternal mortality rates are one of the most significant health inequities amongst all conventional population perinatal health measures within the Unites States (U.S.) (1). Black women in particular have a pregnancy-related mortality ratio three to four times higher than White women (2). Cardiovascular conditions such as cardiomyopathy, preeclampsia and eclampsia are major contributors to maternal mortality in which Black women are disproportionately affected (3). Despite these alarming trends, 80% of pregnancy-related deaths and severe maternal morbidities in the U.S. are said to be preventable (4). Research shows 60 to 75% of excess mortality amongst Black women could be prevented with increased paternal involvement (5). In 2010, The Commission on Paternal Involvement in Pregnancy Outcomes strongly recommended the full inclusion of fathers in pregnancy and family healthcare to reduce and eliminate racial/ethnic disparities in birthing outcomes across the U.S (The Commission on Paternal Involvement in Pregnancy Outcomes 2010). A recent Centers for Disease Control and Prevention (CDC) review on the comprehensive analysis of maternal deaths across nine states by the Maternal Mortality Review Committees (MMRC) found that the most common root causes were patient and family factors (e.g. limited knowledge of warning signs and when to seek care, socioeconomic environment, and chronic disease) (3). However, provider, facility and health system factors (e.g. inadequate training, missed or delayed diagnoses, poor communication, and lack of coordination between clinicians) had a preponderance of detrimental impact (3, 6). It is also important to consider patient and family factors such as limited knowledge of warning signs and when to seek care are resultant of barriers to health education posed by structural inequalities within the healthcare infrastructure (7). Furthermore, there is an evidence gap on the role of racism (i.e. implicit bias) as a psychosocial stressor which negatively impacts perinatal health outcomes for Black women (8, 9). These alarming trends and persistent inequities by race/ethnicity and geographic location reinforce the need to focus on ensuring quality and safety of maternity care for all women.

Our region of focus is the southeastern U.S. Black maternal health care problems are pervasive in the South — particularly in Georgia and South Carolina, two states with particularly high Black maternal mortality rates (6, 10).5,8 Georgia’s pregnancy-related mortality ratio is among the highest in the U.S., exceeding the national figure and the Healthy People 2020 goal by fivefold (11). Almost half of Georgia’s 159 counties are destitute of a maternity provider; in addition, many rural and peri-urban counties in the state are without a birthing facility. Sadly, rural women in Georgia have a 30% - 50 % greater risk of MM rate than their same race, urban counterparts (6, 10, 12).Almost predictably, rural Black women in Georgia have twice the MM rate of rural White women (10). Georgia’s neighbor, South Carolina, has the ninth worst pregnancy-related mortality rate in the nation (13). In South Carolina, the MM rate is 2.6 times higher for Black and other racially/ethnically minoritized women in comparison to White women (43.3. vs. 16.4 maternal deaths per 100,000 live births, respectively) (13). Rural Black women face a heightened risk of maternal morbidity and mortality due to the shortage of hospitals and doctors, disproportionate rates of poverty and glaring health disparities (2, 14-16).Preventable causes of death that are common amongst rural Black women13 have been linked to health care provider and facility factors such as inadequate staff knowledge, system issues, and difficulties in care coordination (17).

Despite the myriad of personal, social, biological, behavioral, clinical care-, and health system-related factors impacting maternal mortality (MM) and severe maternal morbidity (SMM), there are evidence-based interventions that when deployed consistently and properly, are known to improve the health of mothers, before, during, and after pregnancy. Improvement of both birth outcomes and the women’s health occur when preconception health is optimized. Preconception health focuses on the overall well-being of nonpregnant women and men during their reproductive years (defined here as aged 18–44 years). Most models of preconception counseling (PC) were developed with the primary aim of preventing birth defects (18) .However, PC services should not be limited to clinical care under the umbrella of maternal and child health, but rather quality PC services should also be integrated within primary health care services and other community-based settings (19). Several implementation challenges regarding PC have been noted in the discourse including lack of coverage or provider reimbursement for PC, lack of adequate PC information for providers, absence of PC follow-up systems, and lack of interest in PC (20). Likewise, Kotelchuck (21) identified nine similar issues that need to be addressed when implementing PC: 1) eligibility criteria for women, 2) content of care, 3) timing and frequency of care, 4) provider of care, 5) payor source, 6) motivation to participate, 7) community involvement, 8) public health policy and infrastructure support as well as 9) research and monitoring databases. Thus, additional research is needed to develop implementation strategies for PC prior to testing and application.

Another noteworthy consideration for PC is the misalliance between the participation of fathers in the lives of their partners and children, and their incorporation into the maternal and child healthcare system. Men play an important role and source of support for women during pregnancy, birth and parenting (22). They have demonstrated influence on maternal health-related behaviors such as drinking, smoking, fitness, or nutrition –all of which can shape reproductive outcomes– both before conception and during pregnancy (23). In one study that used Early Childhood Longitudinal Study-Birth Cohort data, men who were more actively engaged with their partner’s pregnancy were more likely to encourage early first trimester prenatal care and reduce cigarette consumption for their partner (24). A 2018 systematic review found that interventions engaging men in maternal and infant health were associated with improved preconception care attendance, skilled birth attendance, facility birth, postpartum care, birth and complications preparedness and maternal nutrition. Despite robust evidence of fathers’ impact on the health of mothers and babies, involving fathers is one of the least explored, articulated, and therefore implemented, aspects of maternal and infant health services.

Research shows that father friendly consultation services can increase their involvement in maternal health services which ultimately results in improved maternal health (25). Father-friendliness refers to the degree in which an organization’s operations encourage father involvement in the services and programs it offers (26, 27). Prior to establishing initiatives for fathers, it is recommended that a father-friendly assessment be used to assess how father involvement is encouraged and identify areas that can be strengthened.

The slow uptake of PC in healthcare and the lack of engagement between women and healthcare providers in shared decision making about preconception health can be attributed to numerous factors including limited resources (lack of time in the clinical encounter, guidelines, and reimbursement); provider barriers (lack of knowledge surrounding preconception health, lack of clarity on specialty responsible for providing preconception counseling, lack of care coordination for preconception health); and patient level factors (lack of knowledge of preconception health, lack of access to care prior to pregnancy) (20, 28-31). Incorporating high quality preconception care with a focus on the implementation of tailored strategies in community-based settings and through a team-based approach of screening, counseling, health behavior modification and treatment in health care settings, and facilitating patient-provider shared decision making in the clinical process has the potential for reducing disparities in maternal and infant outcomes. The development of evidence-based PC programs in clinical settings have been limited by small sample size, single-site recruitment and a lack of frameworks (theoretical or implementation) to support implementation and effectiveness outcomes. Additionally, there is a shortage of research on the implementation of evidence-based PC screening and education tools specifically designed for Black women in community-based settings.

We hypothesize that PC, screening, and self-care training with father involvement will be effective in the prevention and control of cardiovascular conditions that contribute to 3M (22). Therefore, our research team is testing implementation strategies of PC with father involvement, through our regional coalition that consists of Healthy Start programs (see letter of support), community-based organizations (CBOs) and -health centers (CHCs) that predominantly serve Black families. Demonstrating the feasibility, acceptability, and appropriateness of implementing this intervention in community-based settings has the potential to create credibility, enthusiasm and consensus building for scaling up the intervention to Healthy Start programs, CHCs and other community-based settings across the South with high Black maternal mortality and morbidity rates

## 2 Methods

### 2.1 Guiding Frameworks

#### 2.1.1 Community-Based Participatory Research (CBPR)

This project will be conducted through community-engaged research partnerships, or CBPR. A key aspect of CBPR is that the prioritized communities are involved in all stages of the research process. This involvement eases the translation of research to practice, a common barrier in more “traditional” research processes that historically and systemically exclude marginalized communities (32). CBPR methods utilizing high levels of community involvement in the planning process are more likely to result in high levels of benefit, trust, and satisfaction within the population being served, which, in turn, broadens the scope and sustainability of the intervention (33). A key factor in introducing successful programs and interventions to address disparities in maternal mortality and morbidity rates is engaging the affected community (i.e., mothers, fathers, community and clinical providers) from the outset and continuing engagement throughout the planning, development, and implementation of selected interventions (34).

#### 2.1.2 Collective Impact Framework

To conduct well-designed structural interventions and robust evaluations, resources are needed to promote interactions among stakeholders from disparate sectors to plan and develop large-scale meaningful structural interventions that can effectively reduce health disparities in populations and communities. Therefore, this research is also guided by the Collective Impact which is defined as “a disciplined, cross-sector approach to solving complex social and environmental issues on a large scale” (35). Interdisciplinary collaborations are promising approaches to intervening on health disparities. Standardizing approaches for motivating multi-stakeholder collaborations is a critical need in disparities research. This proposed research builds on existing partnerships among sectors in the earliest phases of research design. This early and existing partnerships plans effectively by using a collective impact framework and group facilitation to support consensus building that effectively translates evidence into practice (36). To address the perinatal health disparities in the Georgia maternal mortality and morbidity rates and strengthen the capacity of the maternal/child health workforce, the coalition uses the collective impact framework for addressing perinatal health disparities. This framework meshes well with a CBPR approach in its goal to engage community partners and identify opportunity gaps to ultimately target future intervention and prevention.

### 2.2 Program Description

This pilot study involves two distinct components: (1) a comprehensive needs and assets assessment identifying opportunities and barriers to delivery, uptake and adherence of preconception counseling with father involvement and (2) a pilot preconception counseling program. The pilot preconception counseling program follows a screening, brief intervention, and referral to treatment (SBIRT) model.

#### 2.2.1 Aim 1: Develop theory-informed, evidence-based implementation strategies to address maternal morbidity and mortality

We will conduct a community informed needs and assets assessment with our expanded multi-stakeholder, community-based Perinatal Care Research and Intervention Coalition (PCRIC) to identify opportunities and barriers and develop stakeholder-informed evidence-based solutions for optimal maternal health equity. We will use a socioecological lens to underscore the complex interplay between various levels of the social system as well interactions amongst individuals and their environment within the system. In the needs and assets assessment we will explore women and men’s experiences, integrating their intrapersonal, partner-related, family, community, and socio-cultural contexts to produce one behavioral outcome regarding maternal healthcare-seeking behavior. Mixed-methods data collection approaches will be applied including document review, surveys, and focus groups to better understand MM/SMM health priorities and barriers and facilitators to implementation strategies for preconception counseling and father involvement within our partner communities. This will result in a multifaceted implementation plan that will be developed in concert with our regional coalition.

#### 2.2.2 Aim 2: Conduct a community-informed pilot feasibility study to test the multilevel implementation and uptake of evidence-based cardiovascular risk reduction strategies (Preconception Counseling) for reproductive-aged women, prioritized by the regional coalition needs and assets assessment

We will pilot implementation strategies for the uptake of preconception counseling in women with father involvement using the Reach, Effectiveness, Adoption, Implementation, and Maintenance (RE-AIM) framework (36, 37). We will critically assess the context, identify potential barriers and facilitators, and iteratively adapt the way preconception counseling could be implemented in diverse settings. These findings will support future research implementing scalable and sustainable evidence-based strategies that reduce maternal morbidity and mortality in Georgia and South Carolina’s vulnerable communities. Findings from Aim 1 will be used to adapt the preconception counseling intervention. Additionally, the community identified priorities on the type of evidence-based intervention they want implemented may change. However, our study design to test the implementation strategies in community-based settings will remain the same.

### 2.3 Partners

The pilot will leverage longstanding relationships with the community. We will expand a regionally based, community-focused, multidisciplinary PCRIC to inform priorities and strategies to implement evidence-based interventions on preventable MM and SMM. The PCRIC consists of women, fatherhood involvement programs (e.g., 24/7 Dad, Strong Fathers, Strong Families Coalition), maternity-serving community providers and health systems, as well as Healthy Start coalitions in Georgia and South Carolina as well as their associated Community Action Network(s) (CAN). CANs are comprised of local and state government representatives, early childhood centers, hospitals, community-based advocacy, workforce development agencies, local business, and behavioral health agencies. The PCRIC will lead an integrated approach to understand biological, behavioral, socio-cultural, and structural determinants contributing to Georgia and South Carolina’s high MM/SMM. This project aligns with the priorities of the Center for Maternal Health Equity (CMHE) at Morehouse School of Medicine (MSM), including its focus on community partnerships, interdisciplinarity, training, and dissemination of evidence-based approaches. The overall goals of the PCRIC are to 1) incorporate strategic community partnerships and participation to study and address health disparities in maternal health; and 2) establish an infrastructure to support the development, implementation, and testing of proposed implementation strategies, with a focus on patient-centered evidence-based prevention. The proposed research builds on the existing infrastructure of the PCRIC and the multidisciplinary investigative team’s expertise with cardiovascular disease, perinatal health, implementation science, community-engaged research, social work, father involvement, and social determinants of health research with Black mothers and fathers.

### 2.4 Regional Advisory Board (RAB)

Building a RAB is essential to ensuring that the research amplifies and reflects the target population needs, in accordance with CBPR tenets (37, 38). Researchers using participatory methods have found community input invaluable in the design and adaptation of user-friendly, applicable, and culturally appropriate tools. RAB members will consist primarily of members from our regional coalition: Black mothers and fathers, community partner organizations, community health centers, and health care providers. RAB members will receive introductory training on CBPR and will guide iterative research instrument development and modification, implementation strategies, and the translation of research results. Members of the RAB will receive a $100 gift card for each month of their participation.

RAB members were asked to commit to 6 bimonthly meetings to address specific advisory needs of the study, such as feedback on the intervention, recruitment strategies, and dissemination and translation of results. RAB meetings are to be jointly led by a research team member via Zoom and last approximately 2 hours. Moreover, meetings shall be facilitated using recommended methods for conducting CBPR and community-advisory boards (37).

### 2.5 Needs and Assets Assessment

A community-informed needs and assets assessment will be conducted with our RAB to identify opportunities and barriers to delivery, uptake of, and adherence to preconception counseling with father involvement. The CBPR-driven community-informed needs and assets assessment will highlight and uplift the perspectives, preferences, and priorities of the communities of whom interventions are developed with and for (39). This approach uncovers issues of local relevance that can be applied to interventions addressing complex health issues. Assessment through a CBPR process is the cornerstone for collaborative efforts in facilitating community-driven approaches to advance health equity. The needs and assets assessment entails two separate surveys to be completed by ten CBOs that serve majority Black/African American communities; in addition, to focus groups for community members and healthcare providers. The first survey is a preconception counseling survey, meant to gauge CBO staff background and experience with preconception counseling, if any. The second survey is the Father Friendly Check-Up developed by the National Fatherhood Initiative, meant to gauge the “Father-Friendliness” of the CBOs. The Father Friendly Check-Up contains four “assessment categories”: Leadership Development, Organizational Development, Program Development, and Community Development. The focus groups shall explore Black mothers’ and fathers’ prior experiences and knowledge about preconception health and services, integrating their intrapersonal, partner-related, family, community, and socio-cultural, historical contexts. These discussions will further query the causes and potential solutions to maternal health disparities as well as their recommendations for preconception counseling implementation in their community.

#### 2.5.1 Recruitment and Participants

The RAB will be charged with developing a locally, culturally, and contextually appropriate, noncoercive recruitment and enrollment process. The goal is to create an atmosphere of inclusion and a feeling of importance from taking part; addressing common barriers to enrolling racial/ethnic minorities in research; providing education about preconception counseling; and explaining the results of the research. Ten CBOs located in Georgia and South Carolina were purposively recruited to participate in the needs and assets assessment. CBOs that serve at least 50% Black/African American women, serve women of reproductive age (18-44), have established trust and rapport in the community, and have a focus on providing education and services to women at high risk for adverse perinatal outcomes will be sought out. More specifically, to participate in the preconception counseling and father friendliness surveys via, participants have to be employed at one of the ten CBOs and be able to provide insight into the services provided at their organization. RAB members, the research coordinator and research assistants will promote the survey opportunity by email or by attending CBO meetings/gatherings. Administrators at each CBO will then help to identify 3-4 staff members who will take the surveys. Focus group participants have to be 18 years of age or older, Black/African American, and be currently receiving services at one of the CBOs to participate.

#### 2.5.2 Procedures and Deliverables

Surveys will be administered via Qualtrics. To ensure data collection instruments are comprehended by the community we shall employ a readability assessment tool and have community partner sites in addition to the RAB review all documents prior to implementation. Furthermore, informed consent will be obtained from all focus group participants. Stakeholders who participate in focus groups will receive $50 gift cards plus activation fees. Focus groups will be held over Zoom with a facilitator, cofacilitator, and notetaker with expertise and experience in qualitative data collection methods. A structured guide will be used to review and assess existing survey data as well as reports that describe client/patient satisfaction, reach/coverage, cost, and policies that demonstrate the implementation and integration of site practices. A discussion guide will be developed collaboratively by the research team and RAB members to evaluate the implementation factors described in Table 1 below as well as perceived causes and solutions to Black maternal health disparities.

**Table 1.** Implementation Outcomes and Data Collection Methods

| Implementation Outcome | Working Definition | Measures | Data Collection Methods |
| --- | --- | --- | --- |
| Acceptability | The perception among stakeholders (e.g., consumers, providers, managers, policymakers) that an intervention is agreeable | If mothers, fathers, service providers or support systems found the implementation components agreeable, for example in terms of content or delivery | Focus Groups |
| Adoption | The intention, initial decision, or action to try to employ a new intervention | Percent of and type of service providers and clients (mothers and fathers) who intend to use the intervention | Provider survey |
| Appropriateness | The perceived fit or relevance of the intervention in a particular setting or for a particular target audience (e.g., provider or consumer) or issue | If support systems agree that the implementation strategies are in line with organizational priorities | Provider Survey<br>Focus Groups |
| Feasibility | The extent to which an intervention can be carried out in a particular setting or organization. | If mothers, fathers, service providers and support systems staff agree that the implementation strategies can be successfully undertaken | Focus Groups |
| Fidelity | The degree to which an intervention was implemented as it was designed in an original protocol, plan, or police | Measures such as content, frequency, duration, and coverage as prescribed by its designers. Number and type of adaptations that must be made to implementation strategies including information on how and why | Focus Groups |
| Implementation Cost | The incremental cost of the delivery strategy (e.g., how the services are delivered in a particular setting). The total cost of implementation would also include the cost of the intervention itself. | Cost to deliver the innovation | Document Review Focus Groups |
| Coverage | The degree to which the population that is eligible to benefit from an intervention receives it. | The proportion of support systems staffs' participation in the delivery of the implementation strategy | Document Review Focus Groups |
| Sustainability | The extent to which an intervention is maintained or institutionalized in each setting. | Uptake of implementation strategies by support systems continued at a specified time(s) post the initial intervention | Focus Groups |

Source: (45), (Adapted from (46-48)).

Focus groups will be recorded, and the audio will be transcribed verbatim. The PIs and research assistants will apply an inductive analysis approach to identify key themes that emerge from the data. This data is to be collated according to each implementation outcome. Quantitative data will be analyzed using descriptive statistics (SPSS/SAS). To analyze the father-friendly checkup data we will calculate a score for each assessment category for each site. Each category has a maximum score, so the proportion of the total score in each category will be calculated by dividing the score in each category by the maximum possible score. Together with the RAB, we will identify implementation strategies and factors to include where there are strengths and gaps for each community in regard to preconception counseling. Additionally, we will identify challenges that Black mothers and fathers experience that require additional resources/services. This will result in a multifaceted implementation plan (40, 41) to be developed in concert with our RAB.

### 2.6 Pilot Study

To evaluate the implementation strategies of preconception counseling, we will conduct a pilot study of the intervention to determine the feasibility and acceptability. The main objective of the pilot project is to refine the implementation of evidence-based preconception counseling with father involvement in community-based settings to improve cardiovascular health outcomes before and during pregnancy in Georgia, South Carolina, & Tennessee. To implement preconception counseling, we will be using SBIRT. SBIRT is an evidence-based, comprehensive, integrated, public health approach to the delivery of early intervention and treatment services for persons with substance use disorders, as well as those who are at risk of developing these disorders. MSM has adapted this model for people who are seeking to get pregnant to assess their risks and provide early intervention to at-risk people of reproductive age who want to get pregnant. *Screening* will assess the evidenced-based risks of the individual based on ACOG guidelines and community-identified priority components to identify the appropriate level of education and interventions that can make a positive impact on perinatal outcomes when identified prior to pregnancy. *Brief Intervention* will focus on increasing insight and awareness regarding health behaviors and motivation toward behavioral change. Based on the results of the screening assessment, staff will provide a brief intervention for any topic areas where it was indicated that the participant was at risk for health concerns based on their answers. *Referral to Treatment* will be provided to those identified as needing more extensive treatment with access to reproductive health services and linkage to a clinician.

#### 2.6.1 Site Recruitment and Setting

The recruitment and retention of sites will be based on the successful twenty-year track record of the MSM Community Physicians Network, PCRIC and RAB. Project sites that agree to participate will: 1) become a member of the PCRIC; 2) agree to participate in the pilot; 3) demonstrate capacity to enroll at least 5 patients from vulnerable groups (i.e., Black/African American, 18-44 years old); 4) agree to deliver the intervention; 5) be responsible for the clinical care of enrolled patients; particularly, any hypertensive emergencies detected in which the site can also link women directly to outside sources of care.

We seek to determine whether a specific intervention such as PC implemented through Federal Healthy Starts which are heavily dosed in changing the social environment to induce positive pregnancy outcomes through PC. The goal will be to reduce hypertensive disorders and cardiovascular disease that are associated with 3M while also engaging Black fathers. For this purpose, we have specifically selected Healthy Start programs in Georgia. For this purpose, we have specifically selected Healthy Start programs in Georgia and South Carolina as candidates to pilot test this hypothesis based on the following justifications: (a) Healthy Start is a national program funded by Health Resources and Services Administration (HRSA) with a focus on providing education and services to women at high risk for adverse perinatal outcomes before, during, and after pregnancy (42). Healthy Start programs aim to improve maternal and child health with a focus on reducing the disparities associated with poor birth outcomes. Healthy Start programs play an important role in serving patient populations that might not otherwise be reached through traditional health delivery systems. Healthy Start is an ideal vehicle to deliver effective evidence-based interventions focused on modifiable risk factors. The National Healthy Start Association has tremendous reach, serving some of the nation’s poorest and most at-risk families in 101 communities nationwide. (b) Healthy Start programs are required to facilitate evidence-informed practices to promote healthy weight (42) (c) We will be working with Healthy Start sites serving majority Black/African American communities.

#### 2.6.2 Orientation

The two Healthy Start Sites will be required to attend a virtual orientation session. The objectives of the orientation is to (1) inform sites about the importance of Project IMPACT and (2) discuss the goals of Project IMPACT. Orientation sessions will last between 45 to 75 minutes. All site staff that will be responsible for implementing the intervention are required to attend the orientation.

#### 2.6.3 Participants

Twenty women and men from two Healthy Start sites will be asked to participate. Eligibility criteria for participants included: 1) 18–39 years; 2) self-identify as Black, African American, English speaking, 3) not pregnant at the time of enrollment; 4) receiving services at the partner site.

#### 2.6.4 Intervention

When participants agree to be participants in the intervention, they will receive two preconception counseling intervention sessions. Delivery of the pre-pregnancy counseling intervention will be facilitated by site staff. During the study visits, a site staff member will complete a screening measure verbally with participant that focuses on preconception cardiovascular and lifestyle factors (see Table 2 below for specific measures). The screening measure will prompt the site staff member to complete a brief intervention for factors that the participant is not meeting the recommended guidelines on

**Table 2.** Preconception Screening Tool Assessment Components

| Health Category | Assessment Tool |
| --- | --- |
| Social Determinants of Health<sup>b</sup> | PRAPARE Assessment |
| Family Planning and Contraceptive Behaviors<sup>a</sup> | A modified version of the Reproductive Health National Training Center Client-Centered Reproductive Goals & Counseling Flow Chart |
| General Health<sup>a</sup> | A modified version of the |
| Mental Health<sup>a,b</sup> | GAD-2 (anxiety)<br>PHQ-2 (depression)<br>PC-PTSD-5 (trauma) |
| Substance Use<sup>a,b</sup> | Single self-report question about alcohol use<br>Single self-report question about marijuana use<br>Single self-report question about other drug use |
| Diabetes<sup>a</sup> | A modified version of the prediabetes risk test |
| Genetic History<sup>a</sup> | Single self-report question about knowledge of genetic history |
| Nutrition<sup>a,b</sup> | BFRSS Fruit and Vegetable Intake Questions |
| Physical Activity<sup>a,b</sup> | International Physical Activity Questionnaire, Short Version |
| <sup>a</sup>American College of Obstetrics and Gynecology Recommendation |  |
| <sup>b</sup>Community Recommendation |  |

#### 2.6.5 Deliverables

##### 2.6.5.1 Feasibility Outcome Measures and Analysis

Participants in this study will complete two quantitative surveys (baseline and 2-week follow-up) on preconception health factors and behaviors. The primary feasibility outcome measures are recruitment, consent, and retention in the intervention. The secondary outcomes are demographics and social determinants of health (e.g. age, marital status, education, income) and anthropometric measures (maternal height, weight, and blood pressure which will be taken at predefined intervals). We will also explore the social determinant antecedents of poor maternal and paternal preconception health and health care as we will also be interested in understanding how social current social situation influence primary care health-seeking behaviors and practices. The survey instrument will be developed with the RAB and validated by them prior to administration.

##### 2.6.5.2 Implementation Evaluation Measures and Analysis

We will combine qualitative and quantitative data to understand the extent to which the identified interventions could be successfully implemented in other locations. To achieve this, we will collect data on each RE-AIM dimension as proposed by Kessler et al (43) and Glasgow (44)^67^ Some data will be collected as part of the pilot of the intervention (e.g., preliminary outcomes (effectiveness). Additional data will be collected specifically for the RE-AIM analysis.

To determine intervention *Reach*, we will assess the number of people who participate in the pilot as a proportion of those who are eligible, compare the characteristics of those who do and do not participate, determine if our intended audience benefited from the proposed intervention, and provide detailed information on reach and recruitment issues. We will screen and collect basic contact information (name, phone number, and e-mail address) from all potential participants approached about the intervention. Those who decline participation in the pilot of the intervention will have their screening information permanently de-identified. Screening information will be used to determine the representativeness of those who agree to participate compared to those who decline. We will develop both a web-based survey and interview protocols with input from the RAB. Quantitative data (N=15), using the web-based survey, will be gathered from those who decline about their reasons for choosing to not participate. For those who agree, an interview (N =15) through a phone or Zoom call, will be conducted to determine their perspectives on the proposed intervention, perceived benefits and needs, attitudes/motivation, skills, their behaviors, self-efficacy, and any concerns that might impact their adoption of the intervention.

To evaluate *Effectiveness* (Preliminary efficacy), in addition to the primary outcome, biological, psychosocial, demographic, and program specific parameters will be assessed as potential mediators and moderators of intervention preliminary efficacy. Adoption, Implementation, and Maintenance will be obtained by engaging with diverse key stakeholders. Key informant interviews (KII) (N = 30) will be conducted with providers (N=5), participants (e.g., Black mothers and fathers) (15), community-based organizations (N=5) and community health centers (N=5) to obtain qualitative data across these three RE-AIM dimensions, to determine factors that may enhance organizational adoption and maintenance, and to identify potential adaptations. These stakeholders have been identified because they will either receive the intervention, have ownership of the intervention, host/deliver the intervention, and disseminate or distribute the intervention.

For *Adoption*, we will also examine how the intervention fits with organizational priorities and existing workflow and adoption hesitancy with staff members. Quantitative assessment of implementation will measure intervention responses where appropriate. We will examine intervention delivery. Questions will examine how the intervention was delivered, managed, or enforced. We will track adaptions or changes to the intervention by the diverse settings (i.e. urban vs. rural; Healthy Start vs. CHC), different staff members (e.g., study nurse vs. community health worker), different states (e.g., Georgia vs. South Carolina), and over time. We will examine costs associated with *Implementation* of the intervene and their perceptions on return on investment. Maintenance at the organizational level will be determined by conducting interviews with diverse stakeholders (e.g., leaders at the partner sites, administrative staff, HCPs, CBOs). Examination of factors will include alignment and integration of intervention with organizational mission, objectives, and goals; and how was the intervention institutionalized or normalized int their practice. To evaluate *Maintenance* at the participant level, patient outcomes will be measured at 6 months. Long-term attrition (%) and differential rates by patient characteristics or intervention condition will also be examined.

### 2.7 Dissemination Plan

The findings from this study will be disseminated via academic and community channels. In academic channels, the results will be presented at conferences and published in peer-reviewed journals. In community channels, the results will be presented to our sites, participants, and RAB members. We work closely with our RAB to ensure our findings are accessible to the communities that we serve.

## 3 Discussion

### 3.1 Anticipated Benefits

This pilot study, Project IMPACT, aims to address preconception health needs of Black men and women of reproductive age in the Southeastern United States. We expect that this project will improve both preconception health and maternal health outcomes among participants. In addition, this project will inform community-engaged implementation strategies specifically for interventions tailored for vulnerable communities.

This study will provide necessary and sufficient data to inform and develop a large-scale efficacy trial of PC for women and partners for the prevention of maternal morbidity and mortality. This innovative trial will determine the effectiveness of the intervention in reducing the disparities in cardiovascular risks among those of reproductive age, yielding critical data impacting generalizability and likelihood of implementation of results. Ultimately if successful, this project is an important first step toward advancing maternal health equity and improving perinatal cardiovascular outcomes.

### 3.2 Unique Features

Perinatal care providers are usually siloed by medical specialty (i.e. OB/GYNs vs family practice physicians), by provider discipline (i.e. Doctors of Medicine (MDs) vs advanced practice providers such as Certified Nurse Midwives (CNMs) or Physician Assistants (PAs)), and by training and certification method (medical providers vs community-based practitioners like Doulas). Furthermore: 1) geographic barriers to collaboration between rural, peri-urban, and urban providers; 2) limited interaction between community-based organizations, including those who deliver direct services, and advocacy groups with established health systems; 3) relative lack of inclusion of fathers in the reproductive health education and the development and execution of responsive services as well as 3) lapses in multidirectional communication flow between hospital, consultants, and ambulatory care providers all result in a fragmented and highly variable approach to perinatal care. Thus, leading to suboptimal, disparate care and outcomes. This study creates opportunities for increased linkage to affordable perinatal care via preconception counseling that it tailored for and by the community being served.

This research also has several innovative features: first, prioritizing the engagement of women and fathers, with their families to implement a woman-centered and family responsive PCRIC in Georgia and South Carolina, aligned by practice activity (i.e. care of pregnant and postpartum women), not specialty, certification, or local community. Second, utilizing the tenets of community engagement and community-based participatory research (CBPR) to engage diverse stakeholders and providers, who are serving Georgia and South Carolina’s most vulnerable populations, rather than a traditional provider or research led approach. Third, integrating point of care patient reported outcomes and social determinants, to support HIPAA compliant data sharing and dissemination of best practices. Fourth, leveraging the demonstrated community engagement and community-engaged implementation research expertise of the Morehouse School of Medicine (MSM) as highlighted by its contemporary, cutting-edge leadership with the National COVID-19 Resiliency Network (HHS Office of Minority Health) and the Georgia Community Engagement Alliance (CEAL) Against COVID-19 Disparities Initiative (NIH).

### 3.3 Potential Barriers or Limitations

The potential challenges to implementation and evaluation of implementation strategies may be affected by the COVID-19 pandemic. The research team and community partners may have to adjust in-person outreach and engagement efforts to virtual and other creative formats (i.e., Zoom). Additionally, we anticipate community partners may grapple with changes in funding and staff turnover because of the pandemic. As such, community partners will be expanded to include other potential PCRIC collaborators. Data collection and analysis of implementation and intervention effectiveness outcomes will account for these challenges.

## 4 Conclusion

Should the needs and assets assessment as well as the pilot study yield promising results, our community-informed PC intervention will showcase utility in implementing evidence-based cardiovascular risk reduction strategies during PC for Black adults of reproductive age. Given that the pilot will take place in two majority Black/African American communities served by Healthy Start Sites in Georgia, favorable outcomes can support public health policy making PC more readily accessible. Furthermore, this research could facilitate increased usage of CBPR alongside implementation science to address other maternal health inequities; particularly in the Southeast where Black.

## Data Availability

All data produced in the present study are available upon reasonable request to the authors

## 6 Ethics Statement

All aspects of this program have been approved by the institutional review board of the Morehouse School of Medicine (IRB# 1837865).

## 7 Author Contributions

NHG, LR, Cfare the principal investigators and are responsible for conceptualization of the study. NHG, MH, AM, AH, MF, OTOF, LC, RD, DG, EH, KB, AS, TW contributed to manuscript writing and editing.

## 8 Conflict of Interest

The authors declare that the research was conducted in the absence of any commercial or financial relationships that could be construed as a potential conflict of interest.

## 9 Funding

This research was, in part, funded by the National Institutes of Health (NIH) Agreement OT2HL158287. The views and conclusions contained in this document are those of the authors and should not be interpreted as representing the official policies, either expressed or implied, of the NIH.

## 10 Acknowledgments

The authors would like to thank our coalition of community partners who make this research possible.

## Notes

### Competing Interest Statement

The authors have declared no competing interest.

### Clinical Trial

NCT05987059

### Author Declarations

IRB of Morehouse School of Medicine gave ethical approval for this work.

